# Beyond reductions in depressive symptoms: Functional, social, and household outcomes in a pilot cluster-randomized controlled trial of group interpersonal psychotherapy in rural Uganda

**DOI:** 10.64898/2026.07.16.26358298

**Authors:** Elly Atuhumuza, Jacquellyn Nambi Ssanyu, Roscoe Kasujja, Morris Ndeezi, Shakirah Nakalungi, Crystal Huang, Andrew Fraker

## Abstract

**Introduction:** Group interpersonal psychotherapy may improve outcomes beyond depressive symptoms, but evidence on functioning, social support, and household welfare remains limited. We examined these outcomes using mixed methods in a pilot cluster-randomized controlled trial of a six-week intervention in rural Uganda.

**Methods:** Twenty-four villages were randomized to group interpersonal psychotherapy or enhanced care as usual. Eligible participants were female, aged at least 13 years, with Patient Health Questionnaire-9 scores of 10 or more. Outcomes were assessed at baseline, two weeks, and three months after treatment. Regression models used village-clustered standard errors. Exploratory sensitivity analyses compared controls with 33 intervention participants assessed independently of facilitators and after an honesty and confidentiality prompt. Fourteen interviews and three focus group discussions with 30 intervention participants were analysed thematically and integrated by outcome domain.

**Results:** Of 292 randomized participants, 263 completed the three-month assessment. Intervention participants had lower anxiety scores than controls at three months (mean difference −7.18; p<0.001), higher subjective wellbeing (mean difference 3.70; p<0.001), lower disability scores (mean difference −1.01; p<0.001), greater perceived social support (difference 23.4 percentage points; p<0.001), and more meals reported for children in the previous 24 hours (mean difference 0.62; p<0.001). Household food insecurity was lower in the full-sample analysis (difference −23.3 percentage points; p=0.008), but not in the exploratory sensitivity analysis (difference 1.5 percentage points; p=0.883). Qualitative accounts described recovery as restored capacity to work, manage relationships, care for children, and respond to hardship despite material constraints.

**Conclusions:** These preliminary findings suggest that group interpersonal psychotherapy may improve outcomes beyond depressive symptoms, although household welfare findings were less consistent. Larger trials with independent outcome assessment and longer follow-up are needed.

**Trial registration:** The trial was retrospectively registered with the Pan African Clinical Trials Registry (PACTR202606549854263) on 29 June 2026.

## Introduction

Depression is widely undertreated in low- and middle-income countries, where more than 80% of affected people do not receive adequate care [1,2]. This treatment gap has increased interest in psychological interventions that can be delivered outside specialist mental health services. Group interpersonal psychotherapy (IPT-G), delivered by trained lay workers, has emerged as one of the most scalable responses, with early trials in Uganda [3,4] and recent evaluations of shortened community-delivered formats [5,6] showing consistent reductions in self-reported depressive symptoms. IPT-G is delivered in groups and focuses on interpersonal problems linked to depression, while also creating opportunities for peer support, shared reflection, and practical coping [7]. For this reason, its effects may extend beyond symptom reduction. They may also appear in how women manage relationships, carry out household roles, care for children and return to daily routines disrupted by depression.

Evidence from IPT more broadly shows that benefits can extend beyond depressive symptoms to anxiety, social functioning, relationship quality and perceived social support [8–10]. IPT-G literature, including from Uganda, also indicates effects beyond symptom reduction, particularly for functioning, social support, psychological distress, stigma and social or family relationships [3,4,6,11]. However, findings remain uneven across outcome domains. Qualitative work from rural Uganda has also described perceived changes in productivity, family conflict, community cohesion, and children’s school attendance following IPT-G [12]. However, less is known about whether community-delivered IPT-G is accompanied by measurable changes in household-relevant outcomes such as food security, child feeding and caregiving routines, or how women understand the pathway from improved mental health to everyday household change in settings where poverty and gendered household constraints remain largely unaltered.

This paper examines secondary outcomes from a pilot trial of a six-week community-delivered IPT-G model [13], delivered by StrongMinds, a non-governmental organization implementing lay-worker-led community mental health programs in Africa. The trial was designed to generate preliminary evidence and inform the design, outcome selection, and measurement procedures for a future larger cluster randomized controlled trial. The analysis in this paper is guided by the program theory of change, which links structured group therapy to reduced depressive symptoms through improved coping, attention to interpersonal triggers of depression and support within therapy groups. These proximal changes are expected to contribute to broader improvements in wellbeing, social relationships, daily functioning, labor supply, nutrition and children’s educational outcomes. A companion paper reports the primary depression outcome and examines the potential influence of assessment conditions on self-reported outcomes [13]. The present paper focuses on whether reductions in depressive symptoms are accompanied by changes in selected secondary outcomes.

Using a mixed-methods design, we examine mental health and wellbeing, functional capacity, perceived social support, and household welfare, including child-feeding indicators. We integrate qualitative data to examine how women understood these changes and the pathways through which improved mental health was, or was not, translated into everyday household life. Because the companion paper showed that depressive symptom estimates were sensitive to assessment conditions, this paper also examines whether selected secondary outcomes showed similar sensitivity to assessment context. These analyses are treated as exploratory sensitivity analyses. In doing so, the paper contributes to evidence on the broader effects of community-delivered IPT-G in low-resource settings, while informing the design of a future larger trial.

## Methods

### Study design and setting

This paper presents a convergent mixed-methods analysis nested within a pilot cluster randomized controlled trial of the StrongMinds six-week IPT-G model conducted in Mayuge District, Eastern Uganda, between June and November 2025. The trial evaluated a community-delivered, lay-worker-led intervention among women with moderate to severe depressive symptoms. Quantitative and qualitative data were collected during the same study period and integrated during analysis and interpretation.

### Intervention and comparator

Participants in the intervention arm received a six-week IPT-G intervention. This is a community-based, manualized adaptation of IPT-G designed for delivery in routine programmatic settings by trained lay therapy facilitators, mostly Village Health Teams (VHTs), and supervised by StrongMinds. Sessions were held weekly, lasted approximately 90 minutes, and were delivered in groups of typically 10 eligible women drawn from the same village. Groups were organized around common interpersonal triggers of depression where possible, including grief, role disputes or disagreements, role transitions or life changes, and interpersonal isolation.

The intervention was structured in three phases to help participants understand depression in relation to their social and relational circumstances, identify interpersonal problem areas, strengthen communication and problem-solving skills, and mobilize support from the group. The initial phase introduced group members, established group norms, provided psychoeducation on depression and its symptoms, reviewed therapy goals, and supported participants to identify their main interpersonal triggers. The middle phase focused on active problem-sharing, peer and facilitator support, solution generation, and practice of new coping and communication strategies. The termination phase reviewed progress, skills gained, changes in depressive symptoms and interpersonal functioning, relapse prevention, and plans for sustaining improvements after the group ended.

Throughout the intervention, facilitators guided participants to share personal difficulties, reflect on links between interpersonal stressors and depressive symptoms, develop practical responses, and apply these strategies between sessions through weekly homework. Delivery was supported by facilitator training, supervision, and quality monitoring to promote fidelity to the intervention model.

Participants assigned to the control arm received enhanced care as usual. This included a one-time psychoeducation session on depression, screening using the PHQ-9 and referral to the nearest public health facility or psychiatric nurse for further evaluation and management. Control participants also received contact information for a psychiatric nurse briefed on the referral protocol. After the final data collection round, they were offered the opportunity to enroll in the six-week IPT-G program.

### Randomization, allocation and blinding

Villages were randomly assigned to either the intervention arm or control. The village was used as the unit of randomization so that all eligible participants within a village received the same treatment status. Randomization was conducted using computer-generated random numbers, with allocation conducted independently from the field research team and concealed until baseline data collection was completed. Participants and intervention providers could not be blinded to allocation because of the nature of the intervention.

### Participants and eligibility

Quantitative trial participants were recruited through the standard StrongMinds community mobilization process. Trained therapy facilitators conducted community sensitization and door-to-door mobilization about depression. Individuals who expressed interest were pre-screened using the Patient Health Questionnaire-4 (PHQ-4), a four-item measure of depressive and anxiety symptoms [14]. Those scoring five or higher were invited to a central screening location where trained research assistants obtained consent to participate and administered the PHQ-9 to confirm eligibility [15]. Eligible participants were female, aged 13 years and above, permanent residents of the study area, and screened positive for moderate to severe depressive symptoms, defined as a PHQ-9 score of 10 or greater. Participants were excluded if they were unable to provide consent, declined or withdrew consent, reported suicidal ideation, or were identified as experiencing gender-based violence requiring immediate referral.

For the qualitative component, participants were purposively selected from women who had received IPT-G. Sampling sought to capture variation in participants’ experiences of therapy, including perceived changes in depressive symptoms, daily functioning, relationships, social support, caregiving, economic activity, and household wellbeing. Participants were also selected to reflect variation in age, group participation, and reported levels of benefit, so that the qualitative data could explore both positive change and the limits of change in participants’ lives.

### Sample size considerations

The pilot cluster randomized trial was designed to generate preliminary effect estimates and inform the design of a future larger evaluation. The sample size was determined pragmatically, based on the number of villages feasible for implementation, the village-level randomization structure and the typical group size for community-delivered group interpersonal psychotherapy. The study covered 24 villages, allowing for a maximum of 12 participants per group per village.

The qualitative data used in this paper were drawn from 14 in-depth interviews (IDIs) and three focus group discussions (FGDs) with therapy participants, with each group comprising about eight to 12 participants.

### Data collection procedures

Quantitative data were collected in-person electronically using KoboToolbox by trained research assistants at three time points: baseline before treatment, two weeks after treatment completion, and three months after treatment completion. Research assistants administered structured questionnaires in each participant’s preferred language, using the same assessment schedule for participants in both study arms.

Research assistants administered the outcome measures independently from the therapy facilitators as far as possible. However, during the early part of the two-week follow-up, therapy facilitators supported participant mobilization to a central location and were visible, though out of earshot, during some interviews. This procedure was changed during the follow-up wave so that subsequent interviews were conducted independently of facilitators. In the parent methodological analysis [13], this change was treated as a quasi-experimental protocol variation to assess the potential influence of facilitator presence on self-reported outcomes. A second assessment-context procedure, intended to reduce reporting pressure, was introduced during follow-up data collection: an honesty and confidentiality prompt administered immediately before the outcome measurement modules. Although confidentiality, voluntary participation and compensation were already addressed during consent, the prompt emphasized that the research team was independent from therapy facilitators and StrongMinds program staff, that responses were confidential, that honest reporting was valued whether symptoms had improved or worsened, and that participant compensation was guaranteed regardless of responses. The prompt was randomly assigned at the individual level using a random number generator within KoboToolbox.

The broader qualitative component included semi-structured interviews with supervisors, facilitators, and participants who received therapy. For this paper, the analysis draws primarily on participant interviews and FGDs as the focus is on women’s experiences of functional recovery, social support, caregiving, household routines, and practical agency after IPT-G. Semi-structured interview and FGD guides explored pathways into care, experiences and acceptability of group therapy, barriers and facilitators to participation, and perceived changes in emotional wellbeing, relationships, daily functioning, caregiving, and livelihoods. Interviews and discussions were audio-recorded with consent.

### Outcomes and measures

The parent trial’s primary outcome was depressive symptom severity, measured using the PHQ-9 at two weeks and three months after treatment completion. In this paper, PHQ-9 is reported only to provide clinical context for interpreting secondary outcomes. The present analysis examines selected secondary outcomes that correspond to the proximal pathways in the program theory of change, including mental health and wellbeing, functional capacity, perceived social support and household welfare.

#### Mental health and wellbeing

Anxiety symptoms were measured using the seven-item Generalized Anxiety Disorder scale (GAD-7) [16]. Participants reported how often they had experienced each symptom over the previous two weeks, with responses scored from 0 to 3 and summed to produce a total score from 0 to 21. Higher scores indicate greater anxiety symptom severity. Subjective wellbeing was assessed using the Cantril ladder, a single-item measure of life evaluation in which participants rated their current life on a scale from 0 to 10, with higher scores indicating better perceived wellbeing [17]

#### Functional capacity

Functional capacity was measured using the 12-item WHO Disability Assessment Schedule 2.0 (WHODAS 2.0) [18]. The WHODAS asks about difficulties experienced during the previous 30 days across domains including standing for long periods, household responsibilities, concentration, mobility, self-care, social interaction, maintaining friendships, and day-to-day work. Item responses were averaged to generate a mean WHODAS score ranging from 0 to 4, with higher scores indicating greater functional limitation. We also examined two WHODAS-based indicators: any reported difficulty on one or more WHODAS items, and the number of days in the previous 30 days during which participants were completely unable to carry out usual activities or work because of a health condition.

#### Perceived social support

Perceived social support was assessed using the Multidimensional Scale of Perceived Social Support (MSPSS) [19]. Scores were averaged across items to produce an overall score from 1 to 7, with higher values indicating stronger perceived support. We also examined whether participants reported having someone available to support them when needed.

#### Household welfare

We measured household poverty using the Uganda Poverty Probability Index (PPI), a 10-item scorecard that estimates the likelihood that a household lives below the national poverty line. We used the 2022 Uganda PPI, which is based on the 2020 Uganda National Panel Survey; higher scores indicate a higher probability of poverty [20]. Household food insecurity was assessed using a binary indicator of whether any household member had gone a whole day without eating in the previous two weeks. Among participants with at least one child under 18 years living in the household, we assessed children’s daily welfare using the reported number of meals eaten by children in the previous 24 hours. This was treated as a household-level proxy for children’s daily welfare, not as a direct measure of caregiving quality.

### Patient and public involvement

Patients and members of the public were not formally involved in the design, analysis, interpretation, or reporting of this study. Community members, village leaders, therapy facilitators, and local health workers supported community sensitization, participant mobilization, and implementation of the intervention. Participants also shared their experiences through interviews and focus group discussions, which informed interpretation of the quantitative findings.

### Trial registration

This pilot cluster-randomized trial was retrospectively registered in the Pan African Clinical Trials Registry (PACTR202606549854263) on 29 June 2026. Participant recruitment began in June 2025. Although ethical approval was obtained before recruitment, trial registration was not completed before participant recruitment began. This was an administrative oversight. Nonetheless, the primary outcome and core study procedures were specified in the ethics-approved protocol (Supplementary File S2) before recruitment. Deviations and subsequent refinements to follow-up assessment procedures were reported to the research and ethics committee.

### Ethics approval and consent to participate

The study included procedures to identify and respond to participant distress. Research assistants were trained to observe signs of emotional distress, assess comprehension during informed consent, and refer participants for clinical review where needed. Participants who were severely distressed, suicidal, unable to comprehend study procedures, or identified as experiencing gender-based violence were referred according to the study referral pathway. Referral directories were prepared before implementation, and referred participants were followed up within 48 to 72 hours to confirm access to services and wellbeing. All participants provided written informed consent before participation. For participants aged 18 years and above, written informed consent was obtained directly. For adolescents below 18 years, written assent was obtained from the participant alongside consent from a parent or guardian, in line with the approved study procedures. Emancipated minors, including those who were married, pregnant, parenting, or otherwise living independently, provided written informed consent for themselves as approved by the ethics committee. The consent and assent process was conducted in the participant’s preferred language and covered the study purpose, procedures, voluntary participation, confidentiality, potential risks and benefits, referral options and the right to decline or withdraw without penalty. The study was approved by the Mildmay Research and Ethics Committee (Reference Number MUREC-2025-799 and Uganda National Council of Science and Technology (Reference Number SS3945ES).

### Data analysis

Analyses were done in Stata version 18.5, and followed an intention-to-treat (ITT) approach, with participants analysed according to the village-level assignment of their cluster, regardless of intervention attendance or completion. Baseline characteristics were summarized using means and standard deviations (SDs) for continuous variables and proportions for categorical variables. Baseline comparability between study arms was assessed descriptively using treatment-control differences.

For each outcome, treatment-control differences were estimated separately at two weeks and three months after treatment completion. Continuous outcomes were analysed using linear regression models. Binary outcomes were analysed using linear probability models, with coefficients interpreted as percentage-point differences between study arms. All models used village-clustered robust standard errors to account for correlation among participants within the 24 randomized villages. Inference used the finite-sample correction and cluster-based degrees of freedom implemented by Stata for clustered variance estimates.

Because the parent analysis found that self-reported depressive symptom outcomes were sensitive to assessment conditions, we conducted exploratory assessment-context sensitivity analyses for selected secondary outcomes. Two assessment-context features were considered: facilitator visibility during follow-up mobilization or assessment, and receipt of the honesty and confidentiality prompt. Facilitator visibility was treated as non-randomized because it changed after a mid-fieldwork protocol correction. The honesty/confidentiality prompt was randomly assigned at the individual level. Exploratory sensitivity estimates compared the full control arm with the subgroup of intervention participants assessed without facilitator visibility and after receipt of the honesty/confidentiality prompt. These estimates are interpreted as sensitivity estimates rather than fully randomized intention-to-treat effects.

Results are presented as control means or proportions, full-sample treatment means or proportions, treatment-control differences, 95% confidence intervals, and p-values. The available-case ITT estimates are treated as the main treatment-control estimates. Sensitivity estimates are used to examine whether the pattern of findings was robust to assessment conditions intended to reduce reporting pressure. Analyses used all available observations for each outcome at each assessment. Missing outcome values were not imputed. Participants with missing data for a particular outcome or assessment were excluded only from that outcome-specific analysis. Consequently, analytic sample sizes could vary by outcome and time point.

Qualitative data from interviews and FGDs were transcribed verbatim, translated into English where needed. Data were analyzed thematically using a hybrid inductive-deductive approach. Initial coding was guided by the study’s outcome domains, including mental health and wellbeing, functioning, perceived social support, and household welfare. Within these domains, codes and themes were developed inductively from participants’ accounts to capture how women described perceived changes after IPT-G and the pathways through which these changes occurred. For this paper, qualitative findings were integrated with quantitative results by outcome domain. They were used to contextualize and explain quantitative patterns, particularly around functional recovery, social support, household caregiving, and practical agency.

## Results

### Participant characteristics and baseline comparability

Of 418 individuals assessed, 292 met the study criteria and were enrolled across 24 village clusters: 148 in the IPT-G arm and 144 in enhanced care as usual (Fig 1). Before the three-month assessment, 17 participants allocated to IPT-G and 12 allocated to enhanced care as usual did not complete follow-up. The three-month analytic cohort therefore comprised 263 participants (131 IPT-G and 132 enhanced care as usual).

**Fig 1.**
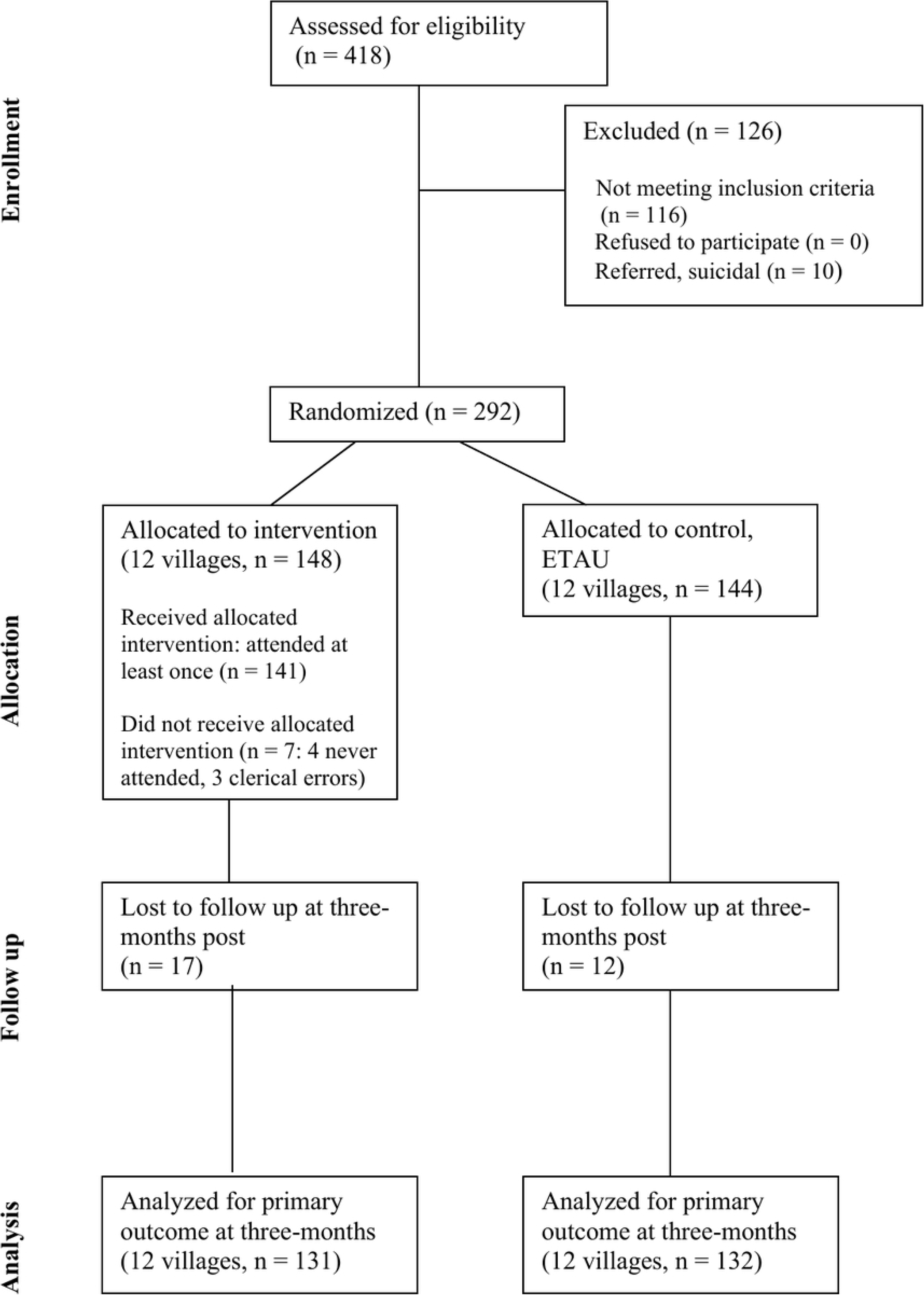
Participant flow through the pilot cluster-randomized trial and three-month quantitative analytic sample. The same randomized cohort is reported in a companion manuscript examining depressive symptoms and follow-up assessment conditions [13].

Baseline characteristics were broadly comparable between randomized arms. Mean age was 33.4 years in the IPT-G arm and 36.6 years in the control arm. Younger participants aged 16-24 years comprised 47.3% of the IPT-G arm and 43.1% of the control arm. The proportion married was slightly lower in the IPT-G arm than in the control arm, but standardized differences were small across measured baseline characteristics. Baseline depressive symptom severity was nearly identical between arms, with mean PHQ-9 scores of 15.62 in the IPT-G arm and 15.60 in the control arm.

Because the exploratory assessment-context sensitivity analysis restricted the intervention group to 33 participants assessed with facilitator independence and after the honesty and confidentiality prompt, we also examined the baseline profile of this subgroup. The subgroup was broadly similar to the control analytic sample on age, marital status, education, number of children, and baseline PHQ-9 score. However, younger participants were somewhat more represented in the sensitivity subgroup, and baseline PHQ-9 scores were slightly lower than in controls. These differences were however not statistically significant (Table 1).

**Table 1:** Baseline characteristics of randomized participants and assessment-context sensitivity subgroup.

| Characteristic | IPT-G randomized (N=148) | Control randomized (N=144) | Standardized difference | p-value | Sensitivity subgroup IPT-G (N=33) | Control analytic sample (N=132) | Standardized difference | p-value |
| --- | --- | --- | --- | --- | --- | --- | --- | --- |
| Age, mean (SD) | 33.38 (15.22) | 36.55 (17.39) | -0.19 | 0.541 | 33.27 (17.55) | 36.55 (17.39) | -0.19 | 0.645 |
| Younger participant, 16-24 years, % | 47.3% | 43.1% | 0.09 | 0.824 | 51.5% | 43.1% | 0.17 | 0.756 |
| Married, % | 41.9% | 50.0% | -0.16 | 0.308 | 42.4% | 50.0% | -0.15 | 0.332 |
| Upper primary education, % | 48.0% | 44.4% | 0.07 | 0.605 | 45.5% | 44.4% | 0.02 | 0.907 |
| Number of children, mean (SD) | 3.59 (2.80) | 3.77 (2.85) | -0.07 | 0.848 | 3.17 (3.05) | 3.77 (2.85) | -0.21 | 0.702 |
| PHQ-9 score, mean (SD) | 15.62 (4.28) | 15.60 (3.92) | 0.00 | 0.983 | 14.58 (4.37) | 15.60 (3.92) | -0.26 | 0.172 |

The qualitative sample comprised 30 female IPT-G participants aged 20 to 57 years, with a mean age of 32.8 years (SD 10.5). Most were married (17/30, 56.7%) or cohabiting (9/30, 30.0%), while three were separated and one was widowed. Educational attainment varied: 14 participants had completed upper primary education, 11 lower secondary education and four lower primary education. One participant had no recorded education level.

### Mental health well-being

The parent trial’s primary depression outcome is reported in the companion paper. Briefly, intervention participants had lower PHQ-9 scores than controls at three months in the available-case ITT analysis (mean difference −7.78 points, 95% CI −9.66 to −5.90; p<0.001). In an exploratory assessment-context sensitivity analysis restricted to intervention participants assessed without facilitator visibility and after the honesty/confidentiality prompt, the estimated difference was smaller but remained clinically meaningful (−5.19 points, 95% CI −7.39 to −2.99; p<0.001).

Anxiety symptoms and subjective wellbeing followed the same pattern. At baseline, GAD-7 and subjective wellbeing scores were similar between study arms (Table 2). In the full-sample analysis, intervention participants reported substantially lower anxiety symptoms than controls at both follow-up points. At two weeks, mean GAD-7 scores were 1.52 in the IPT-G arm and 13.06 in the control arm, corresponding to a mean difference of −11.54 points (95% CI −14.06 to −9.02; p<0.001). At three months, mean GAD-7 scores remained lower in the IPT-G arm than in the control arm (2.51 versus 9.69; mean difference −7.18, 95% CI −8.74 to −5.62; p<0.001). Subjective wellbeing was higher among intervention participants at both follow-up points. At two weeks, mean wellbeing scores were 8.77 in the IPT-G arm and 2.99 in the control arm, a difference of 5.78 points (95% CI 4.67 to 6.88; p<0.001). At three months, the corresponding scores were 7.79 and 4.10, respectively, a difference of 3.70 points (95% CI 2.75 to 4.64; p<0.001).

**Table 2:** Anxiety and subjective wellbeing outcomes: full-sample and exploratory assessment-context sensitivity estimates.

| Outcome and time point | Control (N=132) | Full-sample IPT-G (N=131) | Full-sample difference vs control (95% CI) | p-value | Sensitivity subgroup, IPT-G (N=33) | Sensitivity difference vs control (95% CI) | p-value |
| --- | --- | --- | --- | --- | --- | --- | --- |
| **GAD-7 score** |  |  |  |  |  |  |  |
| Baseline | 12.54 (4.18) | 12.94 (4.43) | 0.40 (-1.05 to 1.86) | 0.594 | 11.24 (4.38) | -1.30 (-3.04 to 0.45) | 0.163 |
| 2 weeks | 13.06 (5.35) | 1.52 (2.97) | -11.54 (-14.06 to -9.02) | **<0.001** | — | — | — |
| 3 months | 9.69 (4.76) | 2.51 (3.07) | -7.18 (-8.74 to -5.62) | **<0.001** | 4.82 (3.58) | -4.87 (-6.57 to -3.17) | **<0.001** |
| **Subjective wellbeing, 0–10** |  |  |  |  |  |  |  |
| Baseline | 2.50 (1.92) | 2.56 (2.01) | 0.06 (-0.55 to 0.68) | 0.838 | 2.39 (1.60) | -0.11 (-0.84 to 0.63) | 0.780 |
| 2 weeks | 2.99 (2.57) | 8.77 (2.04) | 5.78 (4.67 to 6.88) | **<0.001** | — | — | — |
| 3 months | 4.10 (2.42) | 7.79 (2.43) | 3.70 (2.75 to 4.64) | **<0.001** | 6.70 (2.77) | 2.60 (1.13 to 4.07) | **0.003** |

In the exploratory assessment-context sensitivity analysis, the estimated three-month differences were smaller but remained evident for both outcomes. Mean GAD-7 scores were 4.82 in the sensitivity subgroup and 9.69 in the control group, a difference of −4.87 points (95% CI −6.57 to −3.17; p<0.001). Mean subjective wellbeing scores were 6.70 and 4.10, respectively, a difference of 2.60 points (95% CI 1.13 to 4.07; p=0.003).

Qualitative accounts suggest that improvements in anxiety and subjective wellbeing were experienced as a reduction in persistent *“thoughts”* that had previously disrupted sleep, appetite, social interaction and self-worth. Participants commonly described depression through local phrases of *“too many thoughts”, “no peace”,* or feeling unable to continue with ordinary life. After IPT-G, women described feeling calmer, sleeping better, eating with appetite, and being less consumed by distress.

> *“I used to eat food, but it was tasteless. Right now, I eat very well. I used to walk with a broken heart, which is not the case right now. Every time I could sleep, I would hear everything that is outside, right now I sleep very well.”* - IDI Participant 04

### Functional recovery

At baseline, the prevalence of any functional difficulty was similarly high in both study arms (Table 3). In the full-sample analysis, intervention participants reported better functional outcomes than controls at both follow-up points, with larger differences at two weeks than at three months. At three months, 37.4% of intervention participants reported any functional difficulty compared with 88.6% of controls, a difference of −51.2 percentage points (95% CI −62.2 to −40.3; p<0.001). Mean WHODAS scores were also lower in the intervention arm than in the control arm (0.55 versus 1.56; mean difference −1.01, 95% CI −1.31 to −0.72; p<0.001). Intervention participants reported fewer days completely unable to perform their usual activities during the previous month (1.44 versus 4.67 days; mean difference −3.23, 95% CI −4.48 to −1.98; p<0.001).

**Table 3.**
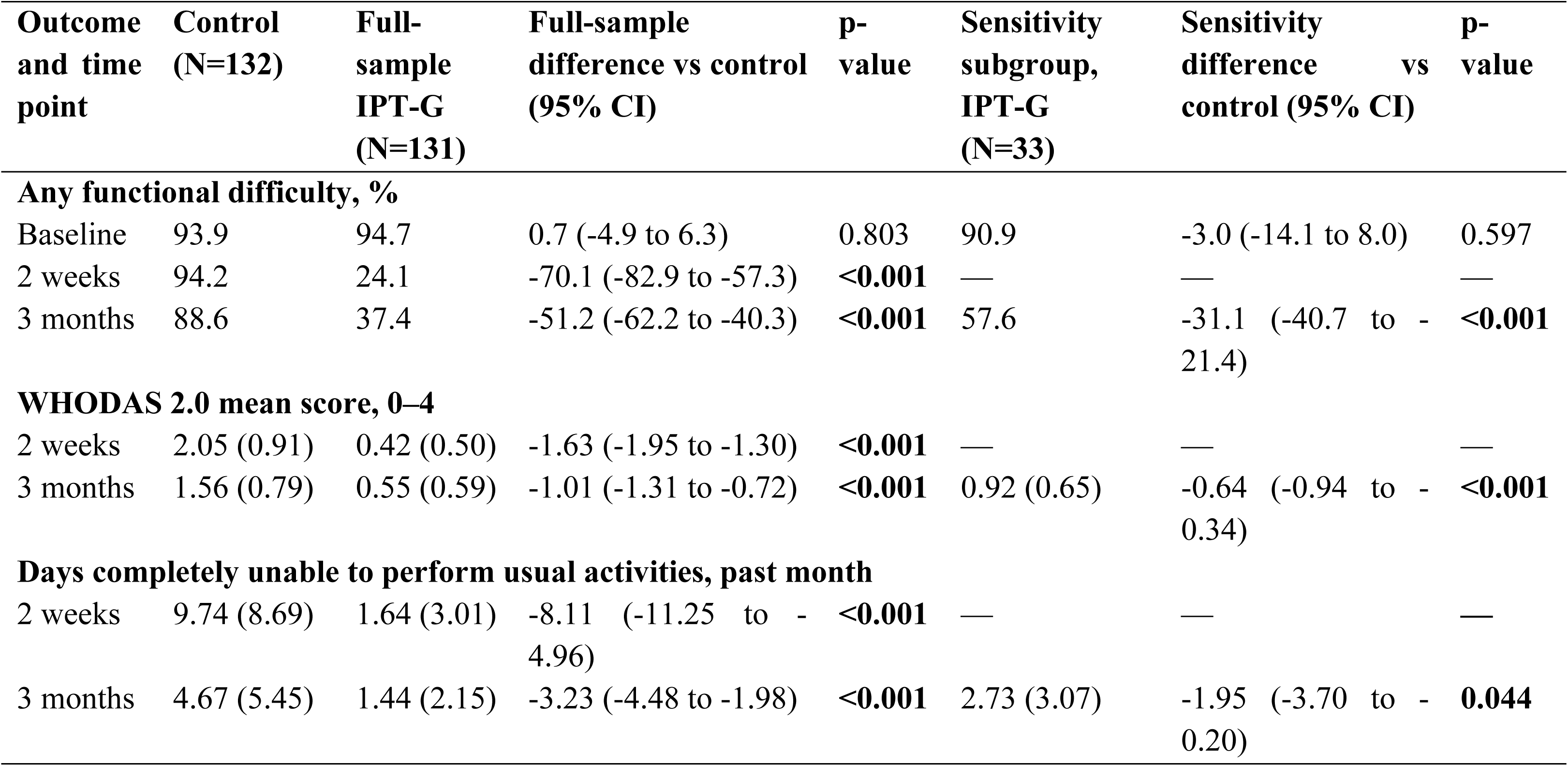
Functional outcomes: full-sample and exploratory assessment-context sensitivity estimates.

In the exploratory assessment-context sensitivity analysis, the estimated three-month differences were smaller but remained evident across all three functional measures. Any functional difficulty was reported by 57.6% of participants in the sensitivity subgroup compared with 88.6% of controls, a difference of −31.1 percentage points (95% CI −40.7 to −21.4; p<0.001). Mean WHODAS scores were 0.92 in the sensitivity subgroup and 1.56 among controls, a difference of −0.64 points (95% CI −0.94 to −0.34; p<0.001). Participants in the sensitivity subgroup also reported fewer days completely unable to perform usual activities than controls (2.73 versus 4.67 days; mean difference −1.95, 95% CI −3.70 to - 0.20; p=0.044).

Qualitative accounts helped explain these quantitative findings by characterizing functional recovery as a return of the capacity to act. Before therapy, participants described fatigue, withdrawal from productive activity, and difficulty completing tasks. After IPT-G, they described renewed energy and a return to work, household responsibilities, caregiving, and routine activities. Recovery was therefore experienced not only as reduced distress, but also as a restored ability to participate in everyday life.

> *“Initially I couldn’t even do any work […] right now I can even do casual labor in someone’s garden and get some money to buy soap.”* Participant 2, FGD1

Another participant linked reduced distress directly to routine household activity:

> *“I wake up, go to the garden, come back cook for my children […] Before therapy, I used not to do most of my work […]. Currently, I have nothing bothering my mind and so I do my activities with energy.”* IDI Participant 08

### Perceived social support

Perceived social support was higher among participants assigned to IPT-G than among controls at both follow-up points (Table 4). In the full-sample analysis, 92.9% of intervention participants reported having someone available to support them at two weeks, compared with 53.8% of controls, a difference of 39.1 percentage points (95% CI 24.9 to 53.4; p<0.001). At three months, the corresponding proportions were 90.8% and 67.4%, respectively, a difference of 23.4 percentage points (95% CI 11.9 to 35.0; p<0.001). Mean MSPSS scores showed a similar pattern. At two weeks, mean scores were 5.67 in the IPT-G arm and 4.06 in the control arm, a difference of 1.61 points on the 1 to 7 scale (95% CI 1.09 to 2.13; p<0.001). At three months, mean scores were 5.57 and 4.29, respectively, a difference of 1.28 points (95% CI 0.72 to 1.83; p<0.001).

**Table 4:** Perceived social support outcomes: full-sample and exploratory assessment-context sensitivity estimates.

| Outcome and time point | Control (N=132) | Full-sample IPT-G (N=131) | Full-sample difference vs control (95% CI) | p-value | Sensitivity subgroup, IPT-G (N=33) | Sensitivity difference vs control (95% CI) | p-value |
| --- | --- | --- | --- | --- | --- | --- | --- |
| **Any perceived social support, %** |  |  |  |  |  |  |  |
| 2 weeks | 53.8 | 92.9 | 39.1 (24.9 to 53.4) | <0.001 | — | — | — |
| 3 months | 67.4 | 90.8 | 23.4 (11.9 to 35.0) | <0.001 | 90.9 | 23.5 (11.1 to 35.8) | 0.002 |
| **MSPSS mean score, 1–7** |  |  |  |  |  |  |  |
| 2 weeks | 4.06 (1.59) | 5.67 (1.09) | 1.61 (1.09 to 2.13) | <0.001 | — | — | — |
| 3 months | 4.29 (1.56) | 5.57 (1.22) | 1.28 (0.72 to 1.83) | <0.001 | 5.13 (1.02) | 0.84 (0.29 to 1.38) | 0.008 |

In the exploratory assessment-context sensitivity analysis, the three-month difference in the availability of social support was similar to the full-sample estimate: 90.9% of participants in the sensitivity subgroup reported having support compared with 67.4% of controls, a difference of 23.5 percentage points (95% CI 11.1 to 35.8; p=0.002). The difference in mean MSPSS scores was smaller than in the full-sample analysis but remained present, with scores of 5.13 in the sensitivity subgroup and 4.29 among controls, a difference of 0.84 points (95% CI 0.29 to 1.38; p=0.008).

The qualitative data suggest that these gains reflected more than simply having someone available for support. Participants described the therapy group as a confidential relational space where private distress could be spoken aloud, especially when problems were linked to marriage, co-wives, in-laws, poverty, or family conflict. Several women said they had initially feared that their household problems would become known in the community, but the group’s emphasis on confidentiality made disclosure possible. In one FGD, participants described gaining *“new friends from the group who are able to keep my issues confidential”,* while another participant explained that she had previously carried her difficulties alone but found support once she opened up:

> *“I was alone, going through a difficult situation. When StrongMinds came, I was happy to get new friends I could talk to about my problems and my home. When I opened up, I was helped. Even my facilitator was helpful and caring.”* Participant 4, FGD2

The group also converted disclosure into problem-solving. Participants described sharing problems, receiving advice, comparing their experiences with others, and realizing that they were not alone. They also reported receiving ideas about how to manage conflict, seek help, restart work, save money, or provide for children. As such, IPT-G appeared to create usable support by making distress discussable and linking disclosure to advice and action.

One important form of this usable support was relational coping within the household. Several participants located their distress in marital, co-wife, in-law, and wider family conflict. After therapy, women described managing these relationships differently through communication, emotional regulation, patience, advice-seeking, and reduced escalation. The intervention appeared to strengthen the conditions for social support within the home by helping women communicate differently, regulate conflict, and rebuild strained relationships.

### Household welfare

Household material and child-feeding outcomes showed a more mixed pattern than the mental health, functioning, and social support outcomes (Table 5). In the full-sample analysis, the PPI-estimated probability of living below the national poverty line was lower in the IPT-G arm than in the control arm at both follow-up points. At two weeks, mean probabilities were 0.16 in the IPT-G arm and 0.30 in the control arm, a difference of −0.14 (95% CI −0.21 to −0.08; p<0.001). At three months, the corresponding values were 0.17 and 0.29, respectively, a difference of −0.11 (95% CI −0.18 to −0.05; p=0.002). In the exploratory assessment-context sensitivity analysis, the three-month estimate was similar: 0.17 in the sensitivity subgroup compared with 0.29 among controls, a difference of −0.12 (95% CI −0.21 to −0.02; p=0.026).

**Table 5:** Household material conditions and child-feeding outcomes: full-sample and exploratory assessment-context sensitivity estimates.

| Outcome and time point | Control (N=132) | Full-sample IPT-G (N=131) | Full-sample difference vs control (95% CI) | p-value | Sensitivity subgroup, IPT-G (N=33) | Sensitivity difference vs control (95% CI) | p-value |
| --- | --- | --- | --- | --- | --- | --- | --- |
| **PPI-estimated probability of living below the national poverty line** |  |  |  |  |  |  |  |
| Baseline | 0.27 (0.22) | 0.29 (0.22) | 0.02 (-0.06 to 0.10) | 0.601 | 0.25 (0.21) | -0.02 (-0.13 to 0.09) | 0.715 |
| 2 weeks | 0.30 (0.22) | 0.16 (0.16) | -0.14 (-0.21 to -0.08) | **<0.001** | — | — | — |
| 3 months | 0.29 (0.22) | 0.17 (0.18) | -0.11 (-0.18 to -0.05) | **0.002** | 0.17 (0.18) | -0.12 (-0.21 to -0.02) | **0.026** |
| **Household went a whole day without eating in the past two weeks, %** |  |  |  |  |  |  |  |
| 2 weeks | 62.0 | 12.1 | -49.9 (-67.8 to -32.0) | **<0.001** | — | — | — |
| 3 months | 43.9 | 20.6 | -23.3 (-39.1 to -7.6) | **0.008** | 45.5 | 1.5 (-18.4 to 21.5) | 0.883 |
| **Children's meals in the previous 24 hours** |  |  |  |  |  |  |  |
| Baseline | 2.02 (0.78) | 1.74 (0.87) | -0.28 (-0.58 to 0.01) | 0.071 | 1.93 (0.77) | -0.10 (-0.44 to 0.25) | 0.593 |
| 2 weeks | 1.66 (0.77) | 2.55 (0.74) | 0.88 (0.59 to 1.18) | **<0.001** | — | — | — |
| 3 months | 1.84 (0.79) | 2.45 (0.77) | 0.62 (0.32 to 0.91) | **<0.001** | 2.45 (0.69) | 0.61 (0.25 to 0.97) | **0.005** |

Reported food insecurity also favored the intervention arm in the full-sample analysis. At two weeks, 12.1% of IPT-G participants reported that someone in the household had gone a whole day without eating during the previous two weeks, compared with 62.0% of controls, a difference of −49.9 percentage points (95% CI −67.8 to −32.0; p<0.001). At three months, the corresponding proportions were 20.6% and 43.9%, respectively, a difference of −23.3 percentage points (95% CI −39.1 to −7.6; p=0.008). However, this pattern was not reproduced in the exploratory sensitivity analysis: 45.5% of participants in the sensitivity subgroup reported food insecurity compared with 43.9% of controls, a difference of 1.5 percentage points (95% CI −18.4 to 21.5; p=0.883).

Children’s meal frequency showed a more consistent pattern. In the full-sample analysis, children in IPT-G households were reported to have eaten more meals during the previous 24 hours than children in control households at both follow-up points. At two weeks, mean meal frequency was 2.55 in the IPT-G arm and 1.66 in the control arm, a difference of 0.88 meals (95% CI 0.59 to 1.18; p<0.001). At three months, the corresponding means were 2.45 and 1.84, respectively, a difference of 0.62 meals (95% CI 0.32 to 0.91; p<0.001). The exploratory sensitivity estimate was similar, with mean meal frequency of 2.45 in the sensitivity subgroup and 1.84 among controls, a difference of 0.61 meals (95% CI 0.25 to 0.97; p=0.005).

Qualitative participants described depression as disrupting both practical and emotional caregiving: cooking, feeding children, bathing children, paying school fees, responding patiently, and maintaining household routines. After therapy, women described becoming more able to organize daily tasks, resume work, and use small earnings for children’s food, schooling, and basic needs.

> *“I used to just sit at home without doing anything apart from getting depressed, but currently I am able to do my activities well. The group members gave us ideas on how to do business. […] I started operating my own mukene (small fish) and tomato [vending] business. I used not to do my domestic work like cooking, digging, but currently I do things in time; my children eat in time and every day.”* IDI Participant 08

However, participant accounts also showed that perceived improvement after therapy often coexisted with continuing material hardship. Several participants described entering therapy with expectations of material support, including money, food, business capital, school fees, tailoring machines, or livelihood training. In one focus group, participants said they had expected support for business capital, food such as maize flour or rice, or even housing where lack of shelter was contributing to distress. Although they later described the therapy as helpful, food shortages, school fees, illness, and lack of income remained important sources of stress. At the same time, some participants described using new coping and problem-solving strategies to manage these pressures, including seeking support, borrowing food during periods of shortage, and finding ways to repay later.

> *“I feel good. The things I was thinking of, that almost made me take poison, the facilitator taught me and I let go of them. Sometimes I still get thoughts when I have failed to get what to eat, but I go to the shop and borrow. After that, I look for the money and pay, and I am good.”* Participant 5, FGD 3

## Discussion

This mixed-methods pilot cluster randomized trial examined whether the StrongMinds six-week IPT-G model was associated with changes beyond depressive symptoms. In the available-case ITT analysis, intervention participants reported lower anxiety symptoms, higher subjective wellbeing, better functional capacity, greater perceived social support, lower PPI-estimated poverty probability, lower household food insecurity, and more frequent meals among children than controls. However, the household findings were less consistent than the mental health, functioning, and social support findings. In the exploratory assessment-context sensitivity analysis, differences remained evident for anxiety, subjective wellbeing, functioning, perceived social support, PPI-estimated poverty probability, and children’s meal frequency, whereas the difference in household food insecurity was no longer observed. Qualitative accounts suggest that participants experienced recovery not simply as reduced distress, but as a restored capacity to act: to resume daily activities, manage relationships, care for children, and respond to continuing material hardship.

The mental health and wellbeing findings suggest that changes following IPT-G were not confined to depressive symptoms. Intervention participants also reported lower anxiety and higher subjective wellbeing, consistent with previous evidence that interpersonal psychotherapy may improve broader psychological distress and wellbeing [3,5,6]. Qualitative accounts provided a corresponding description of this change. Women spoke of relief from persistent “thoughts,” improved sleep and appetite, and a greater sense of calm. These accounts indicate that emotional recovery was experienced in cognitive, physical, and social terms rather than only as a reduction in a symptom score. This is important because it shows that the wellbeing gains captured quantitatively were not abstract scores alone, but reflected changes in how participants felt, slept, ate, and carried themselves through daily life.

The findings on functional capacity extend evidence that improvement in depressive symptoms may support recovery in ways that matter directly for daily survival and household management through reduced disability and increased daily functioning [1,21]. Intervention participants reported fewer functional difficulties, lower WHODAS scores, and fewer days unable to carry out usual activities. Although these effects were smaller exploratory assessment-context sensitivity analysis, they remained statistically significant. Qualitative accounts described this change as a return of capacity, as women and girls who had previously withdrawn from work, household responsibilities, or caregiving described resuming gardening, casual labor, trade, cooking, cleaning, and caring for children. Functional recovery thus emerges as a central pathway through which psychological improvement may translate into household-level change. Nevertheless, the present study did not formally test mediation, and the qualitative accounts should therefore be understood as supporting a plausible pathway rather than establishing that reduced depressive symptoms caused subsequent improvements in functioning.

Perceived social support also improved after IPT-G, and the qualitative findings clarify what this support meant in practice. Participants did not describe support only as having someone available; they described the therapy group as a confidential relational space where private distress could be disclosed without fear of exposure. These findings are consistent with broader work on group-based mental health interventions, where recovery is supported not only by the therapeutic content, but also by the connection, trust, and support built within the group [22]. They also point to a further mechanism: the conversion of available support into usable support. Within the group, confidential disclosure was linked to concrete advice and shared problem-solving. This support also appeared to shape how participants managed conflict at home. Rather than removing household strain, IPT-G seemed to give women practical ways to communicate, regulate conflict and draw on relationships that had previously been difficult or unavailable as sources of support. In this way, the intervention strengthened the relational conditions that could support recovery, even where the underlying tensions remained. Improved social support is therefore best understood not as an endpoint in itself but as a pathway through which reduced symptoms were translated into steadier functioning in the relationships that structure women’s everyday lives.

The household welfare findings were more mixed than the other outcome domains. In the available-case analysis, intervention participants had lower PPI-estimated poverty probability, lower reported food insecurity, and higher children’s meal frequency. Children’s meal frequency showed the most consistent pattern: the estimated difference was similar in the full-sample and exploratory sensitivity analyses. These findings contribute to a smaller and less consolidated literature on the intergenerational effects of maternal mental health treatment [23,24]. In this study, IPT-G appeared to support short-term improvements in children’s daily welfare, particularly through more consistent feeding. However, this should not be interpreted as evidence of longer-term child developmental benefit, as long-term follow-up of a community health worker-delivered perinatal depression intervention in rural Pakistan found no sustained effects on children’s cognitive, socioemotional or physical development at about seven years, despite earlier improvements in maternal depression [25].

The food insecurity result was less robust. Although food insecurity was lower in the intervention arm in the available-case analysis, there was no difference in the exploratory sensitivity analysis. This discrepancy may reflect sensitivity to assessment conditions, differences in the composition of the restricted subgroup, imprecision resulting from the small subgroup, or a combination of these factors. PPI-estimated poverty probability also favoured the intervention arm in both analyses, but this result should be interpreted particularly cautiously. The PPI is a scorecard estimating the probability that a household falls below the national poverty line; it is not a direct measure of income or short-term economic change. A difference in the score over a two-week or three-month period should not be interpreted as demonstrating that IPT-G reduced poverty. At most, the finding indicates that household characteristics incorporated into the PPI differed between arms at follow-up and warrants further investigation using direct and prospectively specified economic measures.

The qualitative findings also suggest caution against interpreting IPT-G as solely a poverty intervention. Participants did not describe therapy as relieving poverty directly. Rather, they described resuming work, drawing on peer advice and small-business ideas from the group, saving small amounts, and using these resources for food, school fees, and other basic household needs. This suggests that the pathway from improved mental health to household change ran through restored capacity to act. Participants were able to resume productive activity and convert small gains in functioning into small but meaningful household resources. This interpretation is consistent with evidence that mental health and economic functioning are reciprocally linked in low-resource settings: improved mental health may support the ability to resume work and generate income, even as food insecurity, illness, school fees, and lack of income remain persistent sources of distress [21,25]. Financial hardship therefore continued to shape how women and girls understood both distress and recovery. The lower PPI-estimated poverty probability alongside continued accounts of hardship is not a contradiction; it suggests that IPT-G may have improved participant’s ability to act within constrained material conditions. The intervention did not remove poverty or livelihood insecurity, but it appeared to reduce some of the psychological and relational barriers that limited women’s capacity to work, plan, seek support and generate small household resources.

Several estimates were smaller in the exploratory assessment-context sensitivity analysis than in the available-case ITT analysis, particularly for anxiety, subjective wellbeing, functioning, and the MSPSS. On the other hand, the food insecurity difference disappeared. This suggests that conventional self-report assessment under standard program conditions in trials of psychosocial interventions for depression may overstate effect magnitudes, not because effects are absent, but because assessment context itself is part of the measured outcome. Participants may report improvement because of gratitude, social obligation, rapport with facilitators, or perceptions of what the program or research team expects. The scale of attenuation observed here is consistent with wider concerns about social desirability in self-reported behavioral and psychosocial outcomes [26]. The exception is also informative. Children’s meal frequency was the only outcome whose magnitude was largely unchanged under the exploratory assessment-context sensitivity analysis. Several explanations are plausible. The measure refers to children’s behavior rather than the participant’s own emotional state, which may reduce the personal stake in the response. It may also be less closely linked to what participants understood therapy to be expected to improve. This implies that the type of outcome being measured shapes how much reporting bias may enter self-reported data.

These findings have implications for the design and evaluation of scalable community mental health interventions. First, they suggest that evaluations of IPT-G should move beyond symptom reduction to include proximal household-relevant outcomes. Second, the attenuation of effects under different assessment conditions underscores the need for independent outcome assessment, strong interviewer training, and protocols that reduce social desirability bias. Third, the poverty-related findings suggest that IPT-G may improve women’s ability to navigate material hardship, even though it does not directly alter the structural conditions that produce poverty. These findings support measuring household economic outcomes in future evaluations and examining whether IPT-G delivered alongside livelihood support produces additive or interactive effects. Finally, because the observed changes were short-term and occurred within persistent poverty and gendered household constraints, larger studies with longer follow-up are needed to assess whether these proximal gains endure and whether they translate into durable improvements in household and child outcomes.

### Strengths and limitations

This study has several strengths. The cluster-randomized design, with allocation concealed until baseline data collection was complete, strengthens causal inference within the limits of a pilot trial, although outcome estimates remain vulnerable to measurement-related bias. A further strength was the study’s direct engagement with measurement bias. The use of two assessment-context variations during follow-up assessment allowed full-sample estimates to be compared with more conservative exploratory assessment-context sensitivity analyses, providing insight into how social desirability may have shaped self-reported outcomes. The mixed-methods design, with integration across outcome domains, also allowed quantitative findings to be interpreted alongside participants’ own accounts of change, rather than against assumed program mechanisms.

However, several limitations should also be considered. First, the assessment-context sensitivity analysis should not be interpreted as a corrected treatment effect. Although the honesty and confidentiality prompt was randomized, facilitator visibility was not; it arose from a mid-fieldwork protocol correction. The sensitivity subgroup was small and may differ from other intervention participants because of timing, village, age composition, baseline severity, or other unmeasured factors. Second, the study was designed as a pilot trial; statistical power and multiple comparisons across secondary outcomes were not specified for definitive inference, and effect estimates should therefore be read as preliminary. Finally, several outcomes were self-reported and may remain subject to reporting bias, including recall error and social desirability, even under the exploratory assessment-context sensitivity analysis.

## Conclusion

In this pilot cluster randomized trial of community-delivered, lay-worker IPT-G in rural Uganda, intervention participants reported lower anxiety, higher subjective wellbeing, better functional capacity, greater perceived social support, and more frequent child feeding than controls at three months post-treatment. Although effect sizes were generally smaller under the exploratory assessment-context sensitivity analysis, the overall pattern suggests that the benefits of IPT-G extended beyond depressive symptom reduction into domains of everyday functioning, relationships, and household life. Qualitative accounts showed that women experienced improvement not simply as feeling better, but as a restored capacity to act: to work, manage relationships, care for children, and respond more effectively to hardship. At the same time, economic insecurity remained central to women’s distress and shaped how they interpreted the limits of the intervention. These findings have implications for the evaluation and design of scalable mental health interventions in low-resource settings, including the importance of measuring household-relevant outcomes and the need to strengthen bias-minimized assessment and examine whether combining IPT-G with livelihood or economic-strengthening components could produce additional gains in household wellbeing.

## Acknowledgements

The authors thank the study participants for sharing their time and experiences. We also acknowledge the research assistants, therapy facilitators, supervisors, community mobilizers, village leaders, and district authorities who supported study implementation and data collection. We thank the StrongMinds Uganda programme and research teams for their contributions to intervention delivery, field coordination, and study oversight.

## Availability of data and materials

The datasets generated and analysed during the current study are not publicly available because they contain sensitive mental health and household information and public release was not covered by the participant consent process. De-identified data and supporting materials may be made available by the corresponding author upon reasonable request, subject to approval by StrongMinds and the relevant ethics and regulatory bodies, and completion of an appropriate data-use agreement.

